# Multi-contrast high-field quality image synthesis for portable low-field MRI using generative adversarial networks and paired data

**DOI:** 10.1101/2023.12.28.23300409

**Authors:** Alfredo Lucas, T. Campbell Arnold, Serhat V. Okar, Chetan Vadali, Karan D. Kawatra, Zheng Ren, Quy Cao, Russell T. Shinohara, Matthew K. Schindler, Kathryn A. Davis, Brian Litt, Daniel S. Reich, Joel M. Stein

## Abstract

**Introduction:** Portable low-field strength (64mT) MRI scanners promise to increase access to neuroimaging for clinical and research purposes, however these devices produce lower quality images compared to high-field scanners. In this study, we developed and evaluated a deep learning architecture to generate high-field quality brain images from low-field inputs using a paired dataset of multiple sclerosis (MS) patients scanned at 64mT and 3T.

**Methods:** A total of 49 MS patients were scanned on portable 64mT and standard 3T scanners at Penn (n=25) or the National Institutes of Health (NIH, n=24) with T1-weighted, T2-weighted and FLAIR acquisitions. Using this paired data, we developed a generative adversarial network (GAN) architecture for low- to high-field image translation (LowGAN). We then evaluated synthesized images with respect to image quality, brain morphometry, and white matter lesions.

**Results:** Synthetic high-field images demonstrated visually superior quality compared to low-field inputs and significantly higher normalized cross-correlation (NCC) to actual high-field images for T1 (p=0.001) and FLAIR (p<0.001) contrasts. LowGAN generally outperformed the current state-of-the-art for low-field volumetrics. For example, thalamic, lateral ventricle, and total cortical volumes in LowGAN outputs did not differ significantly from 3T measurements. Synthetic outputs preserved MS lesions and captured a known inverse relationship between total lesion volume and thalamic volume.

**Conclusions:** LowGAN generates synthetic high-field images with comparable visual and quantitative quality to actual high-field scans. Enhancing portable MRI image quality could add value and boost clinician confidence, enabling wider adoption of this technology.

## Introduction

Magnetic resonance imaging (MRI) has become an essential tool in brain imaging for clinical care and research, but conventional high-field strength MRI scanners (typically 1.5T or 3T) require significant capital investments for siting, operation, safety and staffing. As a result, such MRI units are generally limited to higher level medical, diagnostic and research centers in high income countries^1,2^. These factors restrict access to neuroimaging, including at the point-of-care and in low-resource settings^2,3^.

Recently developed, FDA-cleared, portable, low-field strength MRI scanners operating at 64mT have the potential to increase access to neuroimaging^2,4,5^. These devices are less expensive, more compact, safer, and can be deployed in various clinical and research environments with limited resources and infrastructure. However, the trade-off is less homogenous magnetic fields and lower signal to noise for a given scan time. As a result, low-field images are typically of lower quality and resolution compared to those produced by high-field scanners^4,5^. This limitation may impede adoption and accuracy, particularly for applications demanding higher contrast and spatial resolution, like small lesion detection and volumetrics. For example, portable MRI readily demonstrates white matter (WM) lesions in multiple sclerosis (MS) but tends to miss smaller and more peripheral lesions^6^.

Deep learning techniques have been widely investigated for medical image reconstruction^7,8^, modality transformation^9^, super-resolution^10–13^ and denoising applications^14^. Given the similarity between these prior applications and the signal quality challenges facing low-field MRI, deep learning tools have potential to improve image quality. Portable 64mT brain MRI scanners already incorporate vendor-provided deep learning processing for sequence acceleration and artifact reduction. For super-resolution, Iglesias et al. developed SynthSR, a convolutional neural network trained on synthetic data to generate isotropic 1mm T1-weighted (T1w) MPRAGE-like images from lower quality input images^15^. SynthSR showed impressive results when applied to portable low-field scans for assessing brain morphometry^11^. Taking low-field T1- and T2-weighted (T2w) images as inputs, SynthSR yielded high resolution T1w volumes that strongly correlated with acquired high-field images for multiple brain regions. However, a current limitation of this state-of-the art approach is that it has been designed to generate only T1w outputs and does not provide additional tissue contrasts that may be of clinical interest. Furthermore, because it was not trained on paired data, SynthSR may not capture low-field specific features including pathology present on input images.

In this study, we developed and evaluated a deep learning framework designed to generate high-field quality brain images from low-field inputs. To do so, we curated a unique multi-center paired dataset of MS patients scanned at both 64mT and 3T. As an inflammatory and neurodegenerative disease, MS provides a good model for developing and analyzing our framework, because it causes both WM lesions and brain atrophy. Using paired low-field and high-field scans we developed a multi-contrast generative adversarial network (GAN) for low-to-high-field image translation. We qualitatively and quantitatively compared the synthesized high-field outputs with the actual high-field images. We also compared the performance of our method to that of SynthSR in approximating clinically relevant 3T brain volume measurements from 64mT images, including thalamic volume, an early indicator of neurodegeneration in MS that is correlated with disability and cognitive decline^16,17^. We found that our approach did not artificially generate or obscure white-matter lesions present in the original low-field inputs, preserved total white matter lesion volume, and captured a known inverse relationship between lesion burden and thalamic volume.

## Methods

### Participants

Among adult outpatients undergoing clinical 3T brain MRI for known or suspected MS^18^, 49 were recruited from two medical centers, the Hospital of the University of Pennsylvania (n=25; Age: 46.8±14.2; 20 females) and the National Institutes of Health Clinical Center (n=24; Age: 52.5±12.5; 13 females). The study was approved by the institutional review board (IRB) at the University of Pennsylvania and at the National Institutes of Health, and all participants provided written informed consent.

### MRI Scanning Procedures

All participants received portable 64mT brain MRI scans (Hyperfine SWOOP) on the same day as clinical 3T scans (Siemens) with T1w, T2w, and Fluid-Attenuated Inversion Recovery (FLAIR) sequences at each field strength. High-field scans consisted of 1 mm isotropic 3D T1w and FLAIR images and 0.5 x 0.5 x 5mm or 0.3 x 0.3 x 3mm T2w images. Low-field resolutions were 1.5 x 1.5 x 5mm (T1w and T2w) or 1.6 x 1.6 x 5mm (FLAIR). Additional scanning parameters can be found in Arnold et al^6^.

### Image Preprocessing

To prepare imaging data for model training and testing, we coregistered low-field and high-field scans. All transformations were performed using ANTsPyNet^19^. For each participant, we resampled each low-field and high-field scan into a 1mm isotropic voxel size by linearly interpolating. After resampling, we applied a rigid registration with dense sampling (“dense-rigid”) between each low-field acquisition (T1w, T2w and FLAIR) and the corresponding high-field acquisition for that participant. We additionally computed dense-rigid registrations between high-field T2w and T1w, and FLAIR and T1w. Finally, we transformed every low-field and high-field acquisition into the high-field T1w space. After registration, we skullstripped the high-field T1w image and applied the same skullstripping mask to every other low-field and high-field image. Skullstripping allows for only brain voxels to be included in intensity normalization prior to deep learning. Our final dataset consisted of coregistered 1mm isotropic low-field and high-field T1w, T2w, and FLAIR images for each participant.

### Deep Learning Model

#### Architecture and Training

Low-field and high-field images differ not only in signal-to-noise ratio (SNR) but also in tissue contrast^4^, resulting in fundamental visual differences between low-field and high-field images. For this reason, we consider the problem of going from a low-field scan to a high-field scan as an image translation problem, in addition to a super-resolution problem. To this end, we developed a GAN architecture for low-to-high field image translation which we term LowGAN.

Our proposed architecture consists of two main parts (Figure 1). The first half of the network (stage 1) contains 3 parallel pix2pix layers^20^, each trained using individual low-field slices (2-dimensional) from different orthogonal imaging planes (axial, coronal, sagittal) with T1w, T2w, and FLAIR images as input channels, and the corresponding multichannel slice in the high-field volume as targets. We chose a pix2pix architecture due to its proven versatility in image translation^20,21^, which suits the low-field to high-field transformation. Prior to training each pix2pix network, intensity values for input low-field and target high-field volumes were min-max normalized, and individual 2D slices were reshaped to 256×256 pixels. The generator of each pix2pix layer consisted of a U-Net with the default number of layers and skip connections^20^. Training was done at a fixed learning rate of 1 × 10^-4^ for 30 epochs, followed by a linear learning rate decay for another 70 epochs (100 epochs total), with a batch size of 2. The remaining hyperparameters used were the default parameters in the Pytorch implementation of pix2pix (https://github.com/junyanz/pytorch-CycleGAN-and-pix2pix).

**Figure 1.**
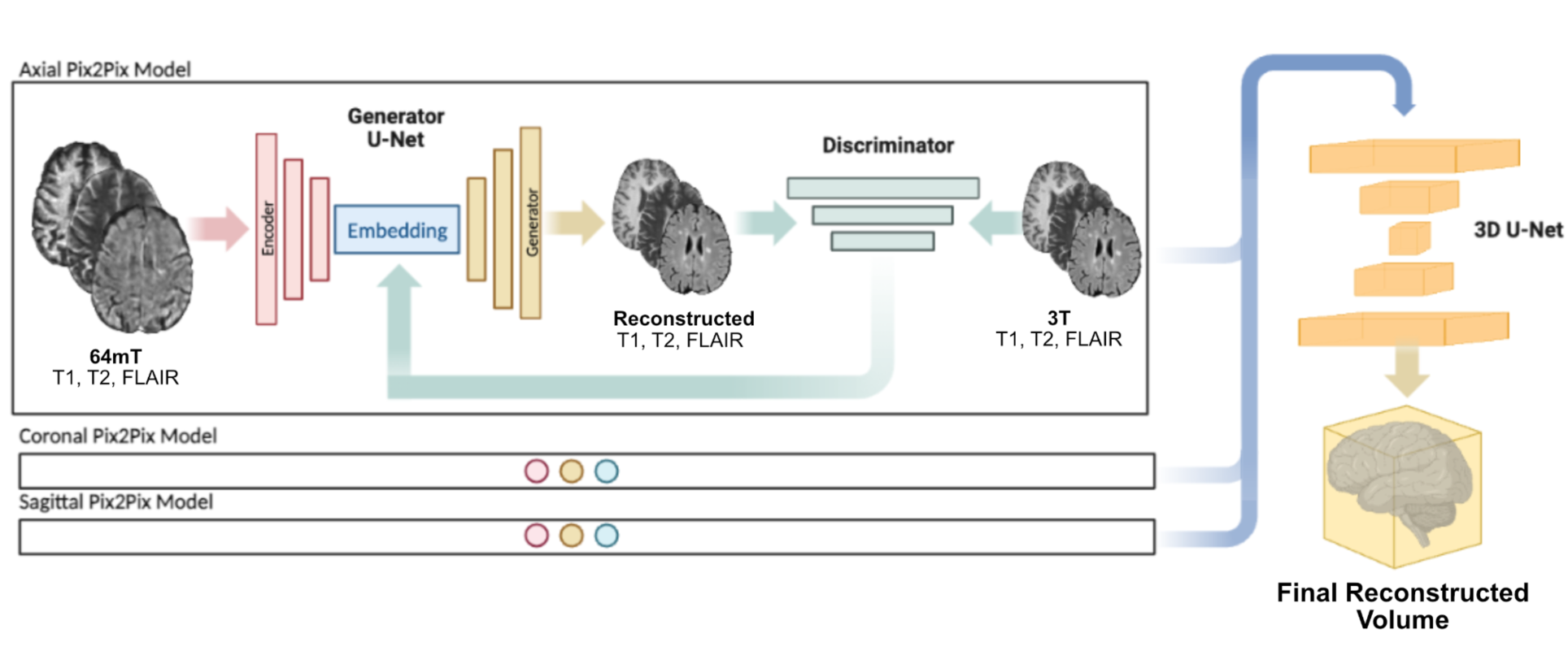
Architecture overview. LowGAN consists of two main stages. Stage 1 (left) consists of 3 parallel pix2pix architectures, each trained on a different orthogonal plane in a 2-dimensional fashion, and using T1w, T2w and FLAIR contrasts as channels. Low-field (64mT) scans are used as inputs to the generator and outputs are compared to paired high-field (3T) scans by the discriminator. Each 2-dimensional output slice is concatenated into a volume which represents the output from stage 1. Stage 2 (right) takes the axial, coronal and sagittal volumes from stage 1 and combines them using a 3-dimensional patch-based U-Net.

The second half of the network (stage 2) consisted of a 3D patch-based U-Net that combines the 3 orthogonal outputs from the first half into a final volume. This step is necessary due to banding artifacts that are generated in the orthogonal planes of the stage one outputs due to intensity fluctuations between adjacent slices (Supplementary Figure 1). The 3D U-Net was trained with reconstructed outputs from the training set of stage 1 as inputs, with each orthogonal plane output (axial, coronal, and sagittal) as a channel, and the corresponding high-field 3D volume as target. T1w, T2w and FLAIR contrasts were used as separate examples for each participant in the same model. Two versions of this second network were trained, one with a mixed L1 with structural similarity (L1-SSIM) cost function, and one with a perceptual cost function for better edge preservation. Results presented in this work are for the L1-SSIM cost function, although our accompanying code outputs for both cost functions.

#### Training and Validation Datasets

We randomly selected 37 participants for training the GAN model. The remaining 12 were used as out-of-sample test participants. Demographics of training and testing groups are shown in Table 1. We evaluated model performance by comparing the normalized cross-correlation (NCC) between acquired low-field and high-field images and the NCC between LowGAN synthetic and high-field images. We chose NCC as a similarity metric because it is capable of measuring how changes in contrast happen across voxels in the two images independent of the actual intensity of the voxels, which might be affected by rescaling and transformation within the neural network.

**Table 1.**
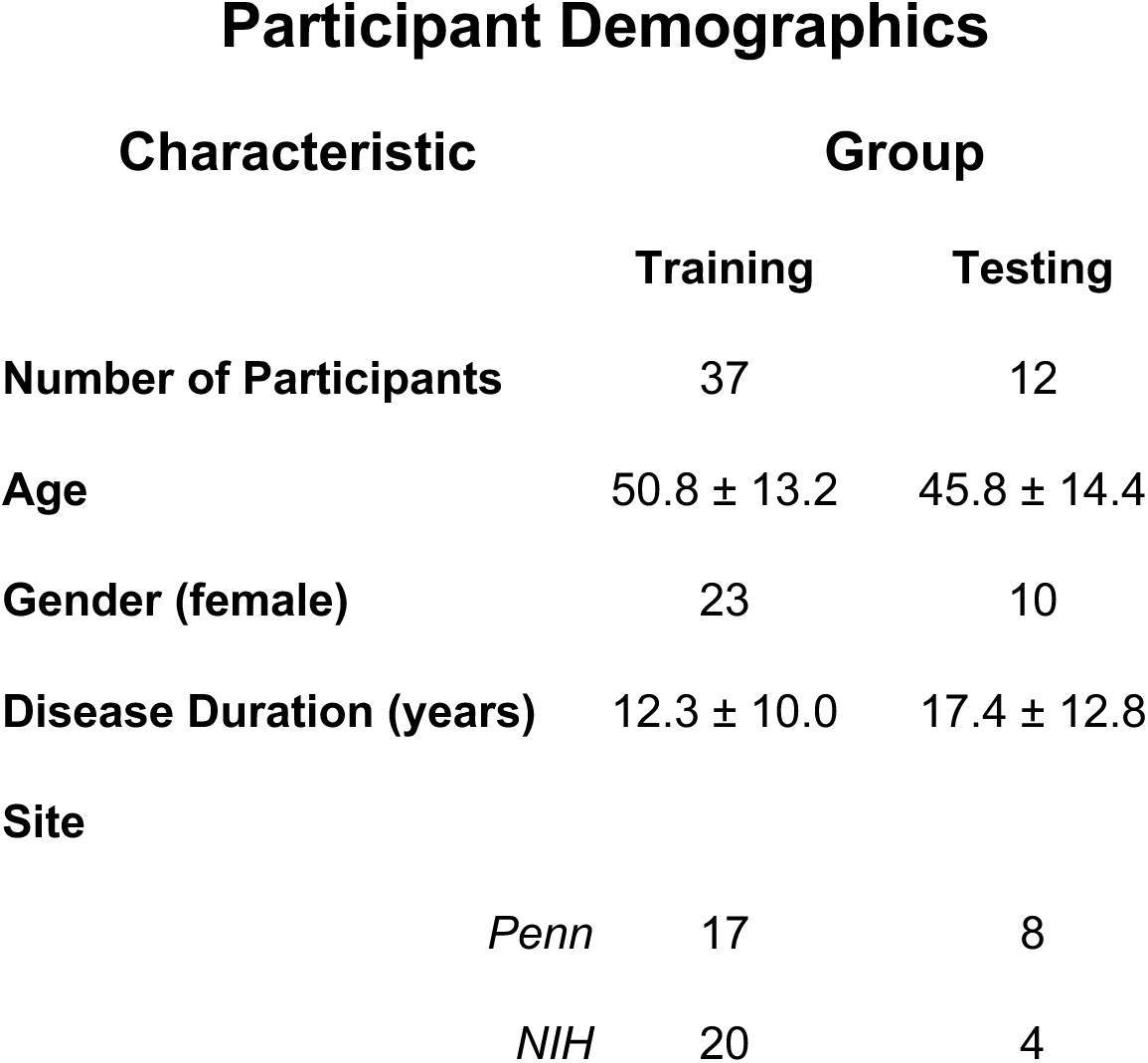
Demographics for training and testing sets.

### Tissue Segmentation and Volumetry

To quantify how LowGAN affected brain tissue contrast and structural feature delineation, we assessed brain tissue segmentation. Because segmentation of raw low-field scans is challenging^22^, we compared segmentations of T1w scans generated by LowGAN with those generated from SynthSR^11^—the current state-of-the-art for low-field super-resolution—and the acquired high-field MRI. To generate SynthSR T1w volumes from low-field images, we used the low-field T1w and T2w sequences of each participant. We generated all segmentations with SynthSeg^23^, a deep learning-based algorithm with robust performance across a range of input resolutions. We then compared various subcortical structures, as well as cortical gray matter, WM, and lateral ventricle absolute volumes between approaches, following similar comparisons performed previously on low-field data by the developers of SynthSR^11^.

### Multiple Sclerosis Lesion Segmentation

We trained and tested LowGAN on participants with MS. However, we did not enforce constraints on the network for it to preserve lesions during the transformation. To assess whether LowGAN could preserve MS lesion burden, we manually segmented lesions in the low-field scans and in the synthetic LowGAN outputs. During segmentation, all three contrasts were available for the low-field scans as well as for the LowGAN outputs. We used MIMoSA^24^, a validated lesion segmentation algorithm, for the high-field scans. We estimated lesion burden as the total volume of segmented lesions in each participant. We then assessed the relationship between total lesion volume and thalamic volume, normalized by intracranial volume.

### Statistical Analyses

We performed all analyses on the outputs of the lowGAN test set. We compared image quality metrics and volumetry between low-field, high-field and LowGAN outputs using pairwise one-sample two-tailed t-tests, with Bonferroni correction for multiple comparisons (pFWER). We used one-sample t-tests since each low-field image had a corresponding high-field image and a corresponding LowGAN output. We used Cohen’s *d* as an estimate of effect size when estimating differences in means, with effect sizes between *d =* 0.5-0.8 considered to be of medium size, and effect sizes *d* > 0.8 considered to be large. To estimate linear relationships between measured volumes across field strengths and LowGAN outputs, we used Pearson’s correlation coefficient. Finally, we estimated the relationship between white matter lesion burden and brain volumes as a decaying exponential fitted using least-squares approximation.

## Results

### LowGAN produces high-field like images

Synthetic high-field output images demonstrated visually superior quality compared to the acquired low-field input images for both stage 1 and stage 2 outputs (Figure 2A-C). This visual enhancement was apparent across T1w, T2w, and FLAIR sequences. In-plane stage 1 outputs visually show the highest improvement relative to low-field inputs, whereas stage 2 outputs provide a smoothed version of the stage 1 outputs. While visually less detailed, stage 2 outputs are necessary for tissue segmentation and downstream processing tasks, as they provide a 3-dimensional volume without banding artifacts (Supplementary Figure 1), which can be successfully utilized by neuroimaging pipelines.

**Figure 2.**
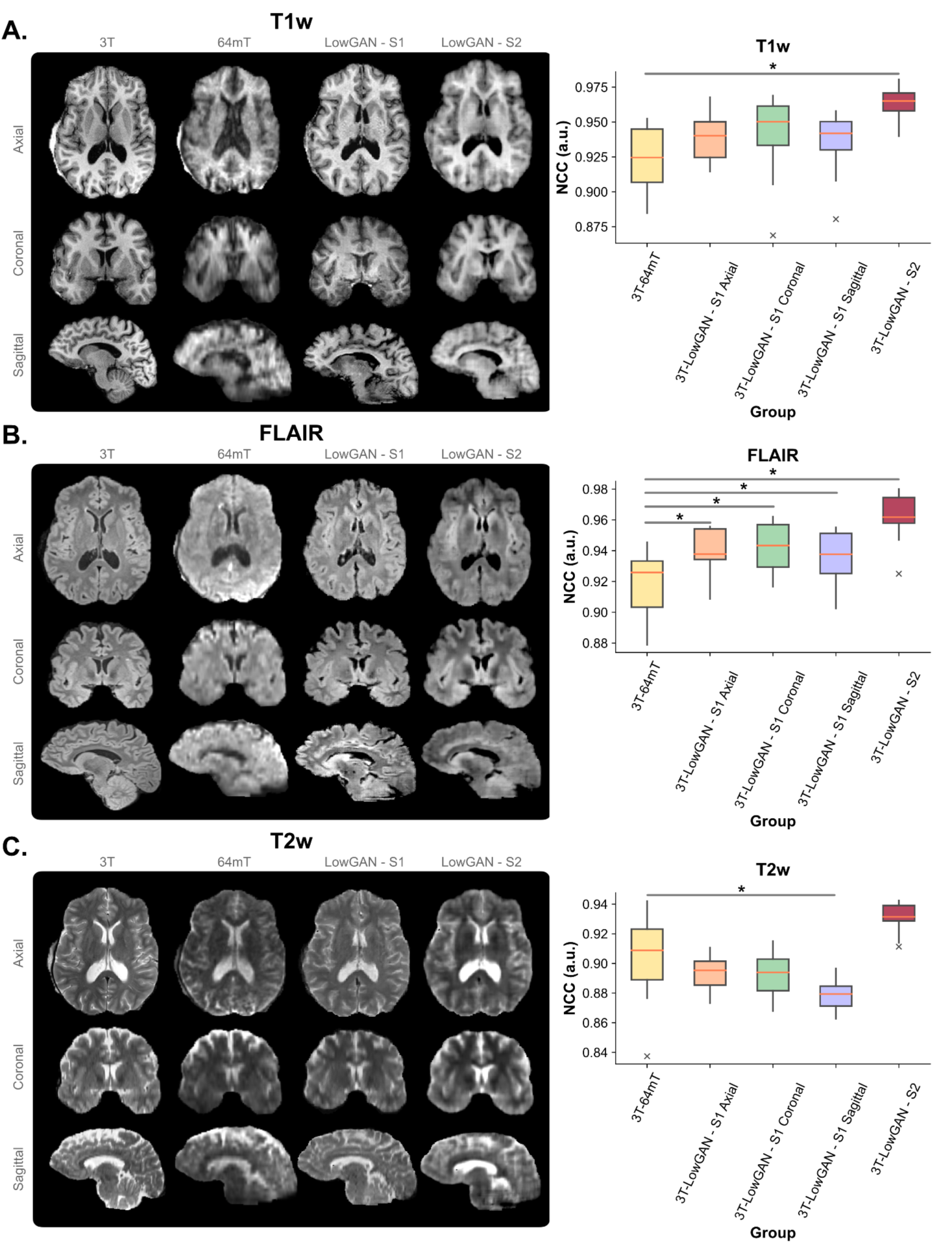
LowGAN improves visual and quantitative image quality. Panels **A**, **B**, and **C** show high-field (3T) and low-field (64mT) images and stage 1 (S1) and stage 2 (S2) LowGAN outputs for a single test set case across T1w (**A**), FLAIR (**B**), and T2w (**C**) contrasts. For stage 1 outputs, each orthogonal view shows the output from the pix2pix model in that plane (e.g. the coronal S1 view is from the coronal pix2pix). To the right of each panel, we show the normalized cross-correlation (NCC) for all cases in the test set between high-field and low-field (3T-64mT), high-field and LowGAN axial S1 output (3T-LowGAN - S1 Axial), high-field and LowGAN coronal S1 output (3T-LowGAN - S1 Coronal), high-field and LowGAN sagittal S1 output (3T-LowGAN - S1 Sagittal), and high-field and LowGAN S2 output (3T-LowGAN - S2). **\*** p<0.05. **a.u.** = arbitrary units.

We quantitatively assessed the quality of the synthetic outputs through the normalized cross-correlation (NCC) between pairs of images. We compared the NCC between LowGAN outputs and high-field images to the NCC between low-field and high-field images (Figure 2A-C and Table 2; 4 comparisons for each output modality). For stage 1 LowGAN FLAIR outputs, we found higher NCC for the axial (p<0.001, Cohen’s *d*=1.26), coronal (p=0.002, Cohen’s *d*=1.29) and sagittal (p=0.014, Cohen’s *d*=0.91) outputs, than for the low-field images. Neither T1w (axial: p=0.37, Cohen’s *d*=2.37; coronal: p=0.63, Cohen’s *d*=0.66; sagittal: p=0.99, Cohen’s *d*=0.51) nor T2w (axial: p=0.65, Cohen’s *d*=0.52; coronal: p=0.65, Cohen’s *d*=0.47) LowGAN stage 1 outputs had significantly higher NCC than corresponding low-field images, and furthermore, sagittal T2w stage outputs had lower NCC (p=0.015, Cohen’s *d*=-1.15) than low-field images. For LowGAN stage outputs, the NCC for T1w (p=0.001, Cohen’s *d*=2.22), and FLAIR (p<0.001, Cohen’s *d*=2.37) outputs was higher than for corresponding low-field images. LowGAN stage 2 T2w outputs also had higher NCC than low-field images, although the difference was not statistically significant after multiple comparison correction (p=0.058, Cohen’s *d*=1.22). Overall, the NCC analysis revealed significantly higher correlation to the paired high-field images for the LowGAN stage 2 outputs in comparison to the input low-field images across T1w, FLAIR and T2w modalities, with the highest improvement in FLAIR, followed by T1w, and then T2w images. These findings suggest that the synthetic images were not only visually superior but also quantitatively more similar to high-field scans.

**Table 2.** Normalized cross-correlation comparison.

| Group | T1w |  |  | FLAIR |  |  | T2w |  |  |
| --- | --- | --- | --- | --- | --- | --- | --- | --- | --- |
| | $\Delta$ NCC (a.u.)<sup>a</sup> | p-value<sup>b</sup> | Cohen's *d* | $\Delta$ NCC (a.u.)<sup>a</sup> | p-value<sup>b</sup> | Cohen's *d* | $\Delta$ NCC (a.u.)<sup>a</sup> | p-value<sup>b</sup> | Cohen's *d* |
| 3T-LowGAN - S1 Axial | 0.016±0.008 | 0.37 | 2.37 | 0.022±0.002 | **< 0.001** | 1.26 | -0.012±0.008 | 0.65 | -0.52 |
| 3T-LowGAN - S1 Coronal | 0.018±0.011 | 0.63 | 0.66 | 0.025±0.005 | **0.002** | 1.29 | -0.011±0.007 | 0.65 | -0.47 |
| 3T-LowGAN - S1 Sagittal | 0.012±0.009 | 0.99 | 0.51 | 0.018±0.005 | **0.014** | 0.91 | -0.026±0.007 | **0.015** | -1.15 |
| 3T-LowGAN - S2 | 0.041±0.008 | **0.001** | 2.22 | 0.044±0.004 | **< 0.001** | 2.37 | 0.027±0.008 | 0.058 | 1.22 |

<sup>a</sup> $\Delta$ NCC is calculated as the difference in NCC between the group and the 3T-64mT NCC (Figure 2)
<sup>b</sup>P-values are Bonferroni-corrected across rows. p-values are for one-sample two-tailed t-tests assessing whether the $\Delta$ NCC is significantly different from zero

**Table 3.**
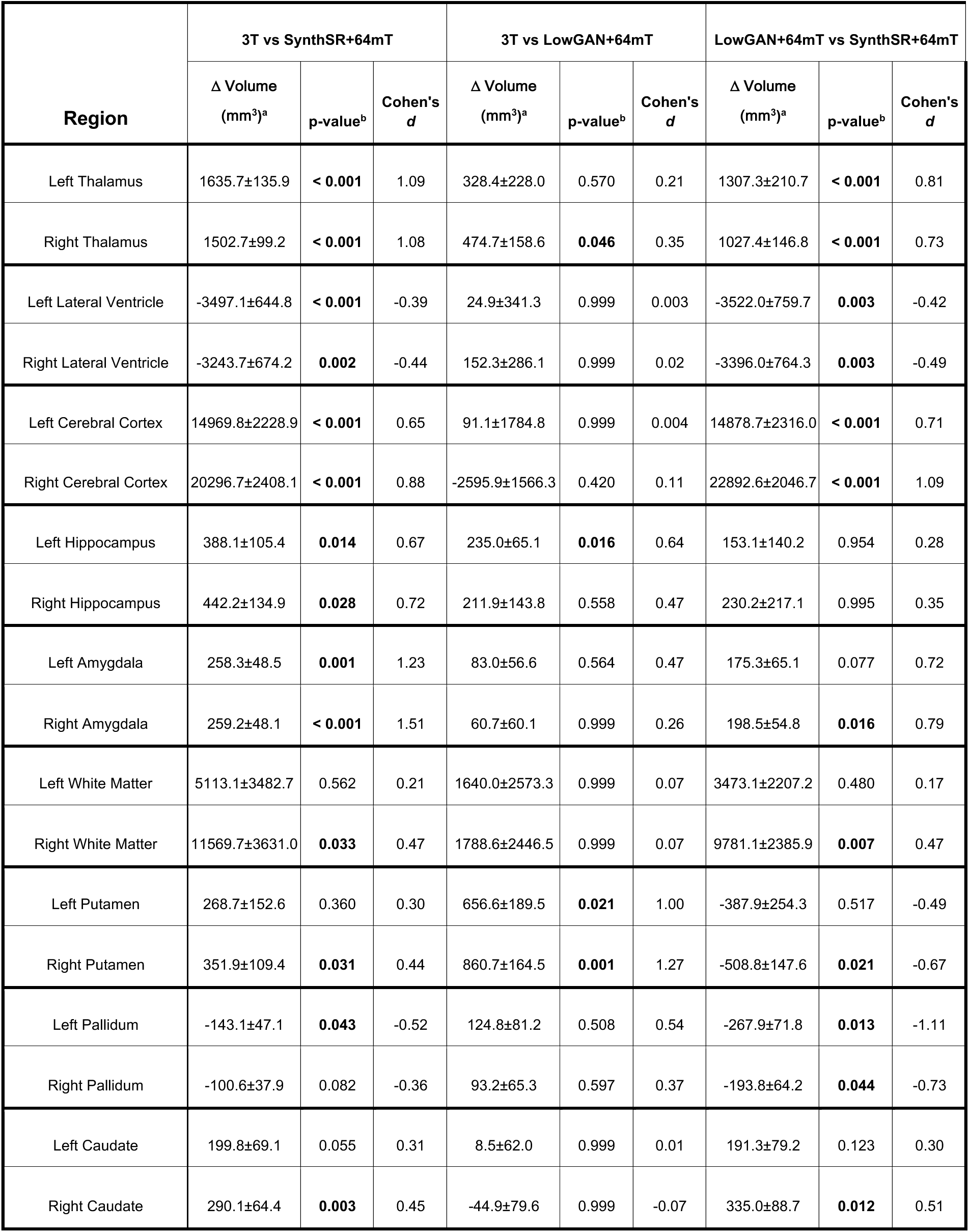
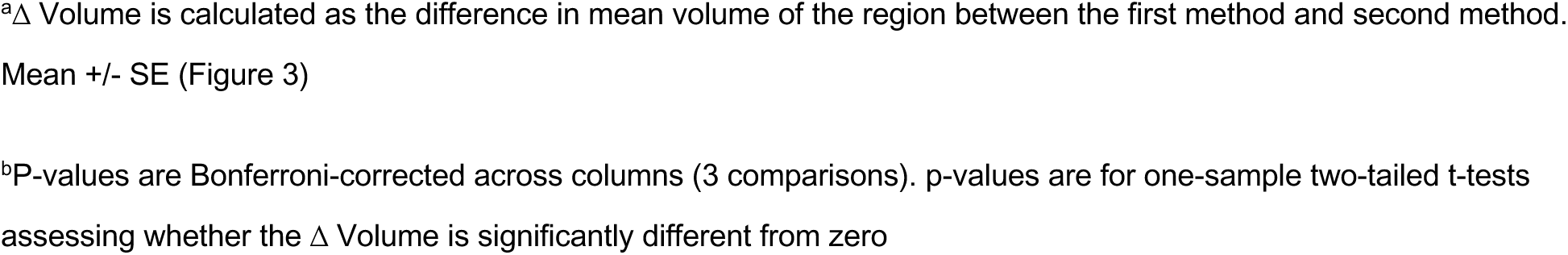
Absolute volumetry comparisons.

| Region | 3T vs SynthSR+64mT |  |  | 3T vs LowGAN+64mT |  |  | LowGAN+64mT vs SynthSR+64mT |  |  |
| --- | --- | --- | --- | --- | --- | --- | --- | --- | --- |
| | $\Delta$ Volume (mm<sup>3</sup>)<sup>a</sup> | p-value<sup>b</sup> | Cohen's *d* | $\Delta$ Volume (mm<sup>3</sup>)<sup>a</sup> | p-value<sup>b</sup> | Cohen's *d* | $\Delta$ Volume (mm<sup>3</sup>)<sup>a</sup> | p-value<sup>b</sup> | Cohen's *d* |
| Left Thalamus | 1635.7±135.9 | < 0.001 | 1.09 | 328.4±228.0 | 0.570 | 0.21 | 1307.3±210.7 | < 0.001 | 0.81 |
| Right Thalamus | 1502.7±99.2 | < 0.001 | 1.08 | 474.7±158.6 | **0.046** | 0.35 | 1027.4±146.8 | < 0.001 | 0.73 |
| Left Lateral Ventricle | -3497.1±644.8 | < 0.001 | -0.39 | 24.9±341.3 | 0.999 | 0.003 | -3522.0±759.7 | **0.003** | -0.42 |
| Right Lateral Ventricle | -3243.7±674.2 | **0.002** | -0.44 | 152.3±286.1 | 0.999 | 0.02 | -3396.0±764.3 | **0.003** | -0.49 |
| Left Cerebral Cortex | 14969.8±2228.9 | < 0.001 | 0.65 | 91.1±1784.8 | 0.999 | 0.004 | 14878.7±2316.0 | < 0.001 | 0.71 |
| Right Cerebral Cortex | 20296.7±2408.1 | < 0.001 | 0.88 | -2595.9±1566.3 | 0.420 | 0.11 | 22892.6±2046.7 | < 0.001 | 1.09 |
| Left Hippocampus | 388.1±105.4 | **0.014** | 0.67 | 235.0±65.1 | **0.016** | 0.64 | 153.1±140.2 | 0.954 | 0.28 |
| Right Hippocampus | 442.2±134.9 | **0.028** | 0.72 | 211.9±143.8 | 0.558 | 0.47 | 230.2±217.1 | 0.995 | 0.35 |
| Left Amygdala | 258.3±48.5 | **0.001** | 1.23 | 83.0±56.6 | 0.564 | 0.47 | 175.3±65.1 | 0.077 | 0.72 |
| Right Amygdala | 259.2±48.1 | < 0.001 | 1.51 | 60.7±60.1 | 0.999 | 0.26 | 198.5±54.8 | **0.016** | 0.79 |
| Left White Matter | 5113.1±3482.7 | 0.562 | 0.21 | 1640.0±2573.3 | 0.999 | 0.07 | 3473.1±2207.2 | 0.480 | 0.17 |
| Right White Matter | 11569.7±3631.0 | **0.033** | 0.47 | 1788.6±2446.5 | 0.999 | 0.07 | 9781.1±2385.9 | **0.007** | 0.47 |
| Left Putamen | 268.7±152.6 | 0.360 | 0.30 | 656.6±189.5 | **0.021** | 1.00 | -387.9±254.3 | 0.517 | -0.49 |
| Right Putamen | 351.9±109.4 | **0.031** | 0.44 | 860.7±164.5 | **0.001** | 1.27 | -508.8±147.6 | **0.021** | -0.67 |
| Left Pallidum | -143.1±47.1 | **0.043** | -0.52 | 124.8±81.2 | 0.508 | 0.54 | -267.9±71.8 | **0.013** | -1.11 |
| Right Pallidum | -100.6±37.9 | 0.082 | -0.36 | 93.2±65.3 | 0.597 | 0.37 | -193.8±64.2 | **0.044** | -0.73 |
| Left Caudate | 199.8±69.1 | 0.055 | 0.31 | 8.5±62.0 | 0.999 | 0.01 | 191.3±79.2 | 0.123 | 0.30 |
| Right Caudate | 290.1±64.4 | **0.003** | 0.45 | -44.9±79.6 | 0.999 | -0.07 | 335.0±88.7 | **0.012** | 0.51 |

<sup>a</sup> $\Delta$ Volume is calculated as the difference in mean volume of the region between the first method and second method.
Mean ± SE (Figure 3)
<sup>b</sup>P-values are Bonferroni-corrected across columns (3 comparisons). p-values are for one-sample two-tailed t-tests assessing whether the $\Delta$ Volume is significantly different from zero

### LowGAN recovers clinically relevant volumes

To quantify how well LowGAN could preserve brain structures, as well as recover clinically relevant volumes, we performed brain tissue segmentation on LowGAN stage 2 outputs using SynthSeg. Example segmentations are shown in Figure 3A. We compared SynthSeg segmentations generated from LowGAN inputs to those generated from SynthSR inputs and high-field inputs by quantifying total volume in subcortical structures, cortical gray matter, lateral ventricles and white matter.

**Figure 3.**
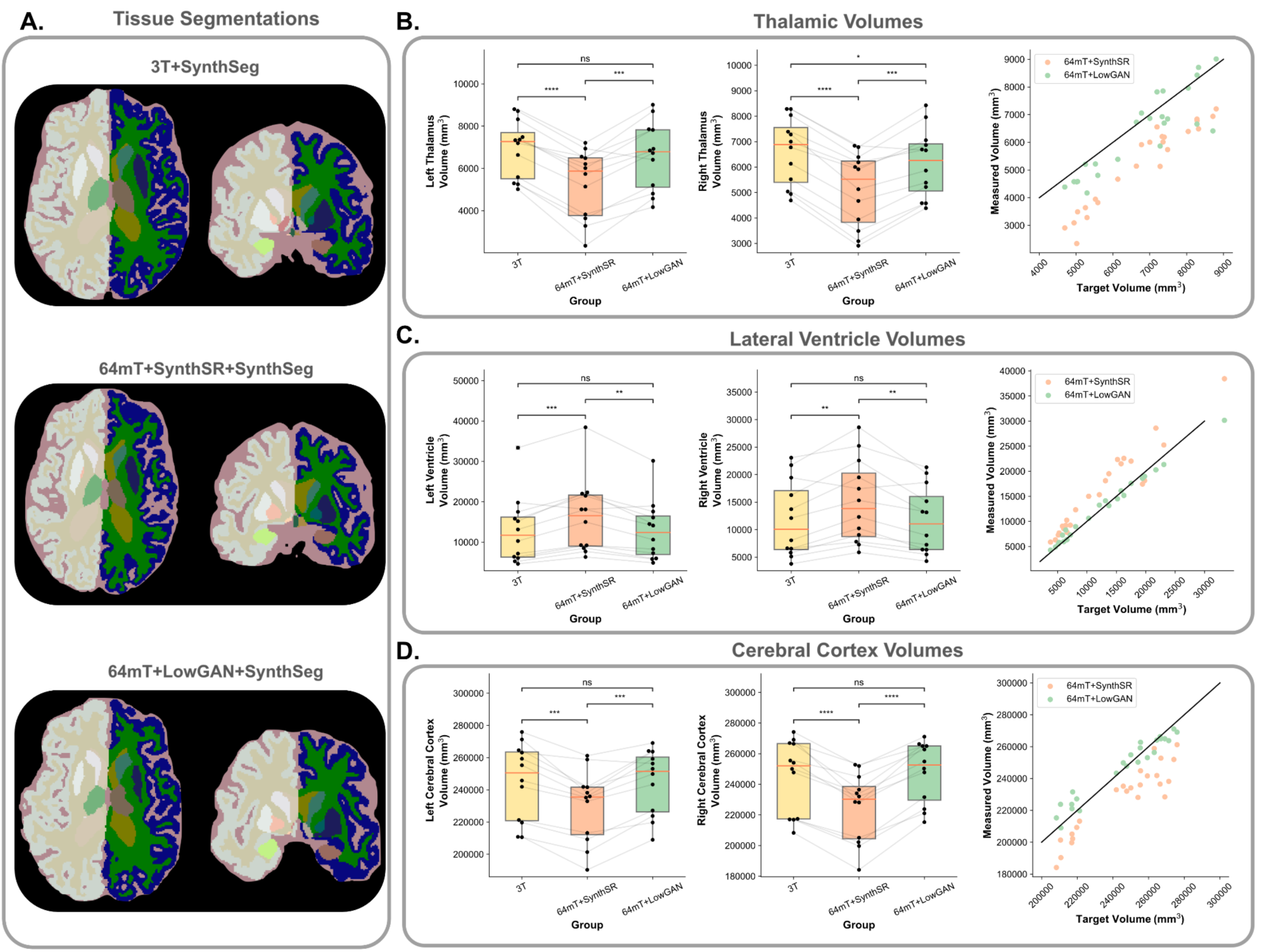
LowGAN outperforms SynthSR for tissue segmentation. Panel **A** shows example SynthSeg segmentations for a single test set case generated from high-field (3T+SynthSeg; top), low-field with SynthSR (64mT+SynthSR+SynthSeg; middle), and low-field with LowGAN (64mT+LowGAN+SynthSeg; bottom) scans. Panels **B**, **C** and **D** show estimated volumes for left and right thalamus (**B**), lateral ventricles (**C**), and cerebral cortex (**D**). Lines connect the same cases across the boxplots for volumes measured at 3T, 64mT reconstructed with SynthSR (64mT+SynthSR), and 64mT reconstructed with LowGAN (64mT+LowGAN). Scatterplots at the right compare the measured volumes for combined left and right sided structures from SynthSR and LowGAN synthesized outputs to the high-field target volumes. The black line represents perfect correspondence between high-field and synthesized volumes.

Absolute volume comparisons across brain structures are reported in Table 2, and total intracranial volume normalized comparisons are reported in Supplementary Table 1. Thalamic (Figure 3B), cortical gray matter (Figure 3D), and amygdala (Supplementary Figure 3) volumes from LowGAN inputs were similar to volumes from high-field inputs, whereas volumes from SynthSR inputs were significantly lower than high-field volumes and LowGAN volumes across these structures. Lateral ventricle volumes (Figure 3C) from LowGAN inputs were similar to lateral ventricle volumes from high-field inputs, whereas lateral ventricle volumes from SynthSR inputs were significantly higher than high-field volumes and LowGAN volumes. In the hippocampus (Supplementary Figure 4), both LowGAN and SynthSR underestimated the structure’s volume, but LowGAN did so to a lesser extent. For basal ganglia structures, LowGAN produces more accurate volume estimates than SynthSR in the caudate (Supplementary Figure 5) and pallidum (Supplementary Figure 7), but not in the putamen (Supplementary Figure 6), where SynthSR outperforms LowGAN. Supplementary Figures 2-8 include Bland-Altman plots for all comparisons.

Overall, LowGAN outputs outperformed SynthSR in producing volumes approximating the high-field standard, correcting undersegmentation in the thalamus and cortex, and oversegmentation in the lateral ventricles. While performance in the hippocampus is still suboptimal, LowGAN has excellent performance in the amygdala. The only subcortical region where LowGAN clearly underperforms is in the putamen.

### LowGAN preserves white matter lesion burden and its relationship with thalamic volume

MS lesions are T2/FLAIR hyperintense and particularly T1 hypointense on the low-field sequences, and we were hopeful that the combination of multiple tissue contrasts would increase lesion conspicuity in the network. Lesions present in the acquired low-field images were preserved in the LowGAN outputs, and in certain cases, made more apparent (Figure 4A). In many cases, lesions were preserved both in stage 1 and stage 2 outputs (Figure 4A), but in other cases stage 1 outputs in certain planes would smooth out the lesions, and only the multiplane stage 2 output would be able to preserve them (Figure 4B). The logarithm of low-field (Pearson’s *r* = 0.95, *p*=1.68×10^-5^) and LowGAN (Pearson’s *r* = 0.91, *p*=2.11×10^-4^) estimated lesion burden were correlated with the logarithm of the high-field lesion burden (Figure 5A,B), although in both cases there was an overestimation of the lesion burden at lower high-field lesion burden. The logarithm of low-field and LowGAN lesion burden were highly correlated (Pearson’s *r* = 0.91, *p*=2.12×10^-4^) and consistent (Figure 5C). Finally, we were able to replicate an inverse relationship between lesion burden and thalamic volume seen at high-field (*r^2^*=0.77) both at low-field (*r^2^*=0.80) and with LowGAN outputs (*r^2^*=0.71), using bilateral thalamic volume estimates from SynthSR and LowGAN respectively. Due to a better estimate of thalamic volumetry with LowGAN, the asymptotic estimate of the relationship (1.06×10^4^ mm^3^, as estimated in high-field), was more accurate for LowGAN (7.61×10^3^ mm^3^) than for low-field (3.90×10^3^ mm^3^).

**Figure 4.**
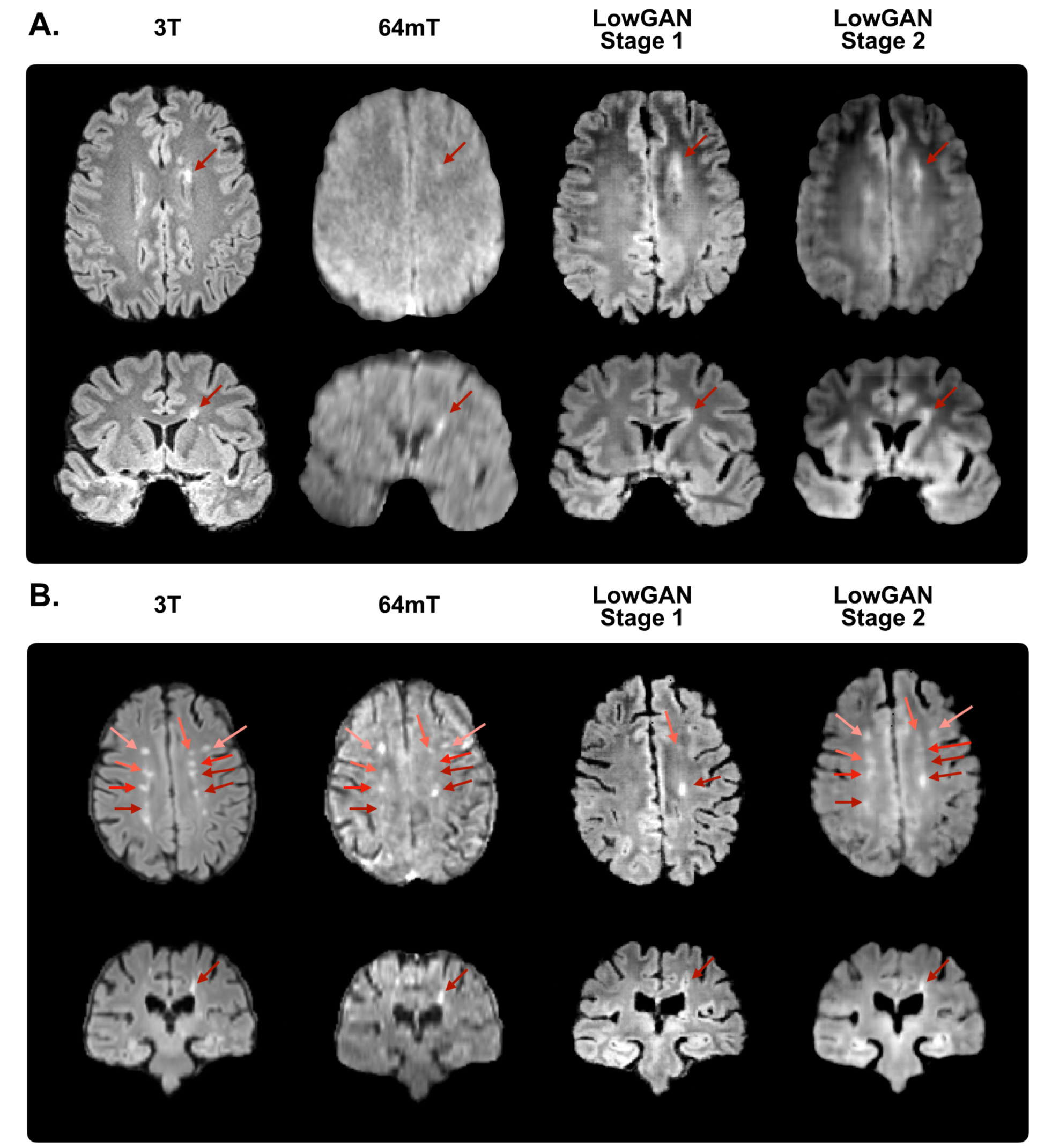
Representative white matter lesions. Panels **A** and **B** show high-field (3T), low-field (64mT), stage 1 and stage 2 LowGAN output FLAIR images for two test set cases with white matter lesions shown by arrows. For stage 1 outputs, each orthogonal view shows the output from the pix2pix model in that plane (e.g. the coronal S1 view is from the coronal pix2pix). The same slice is shown across field strengths/reconstructions.

**Figure 5.**
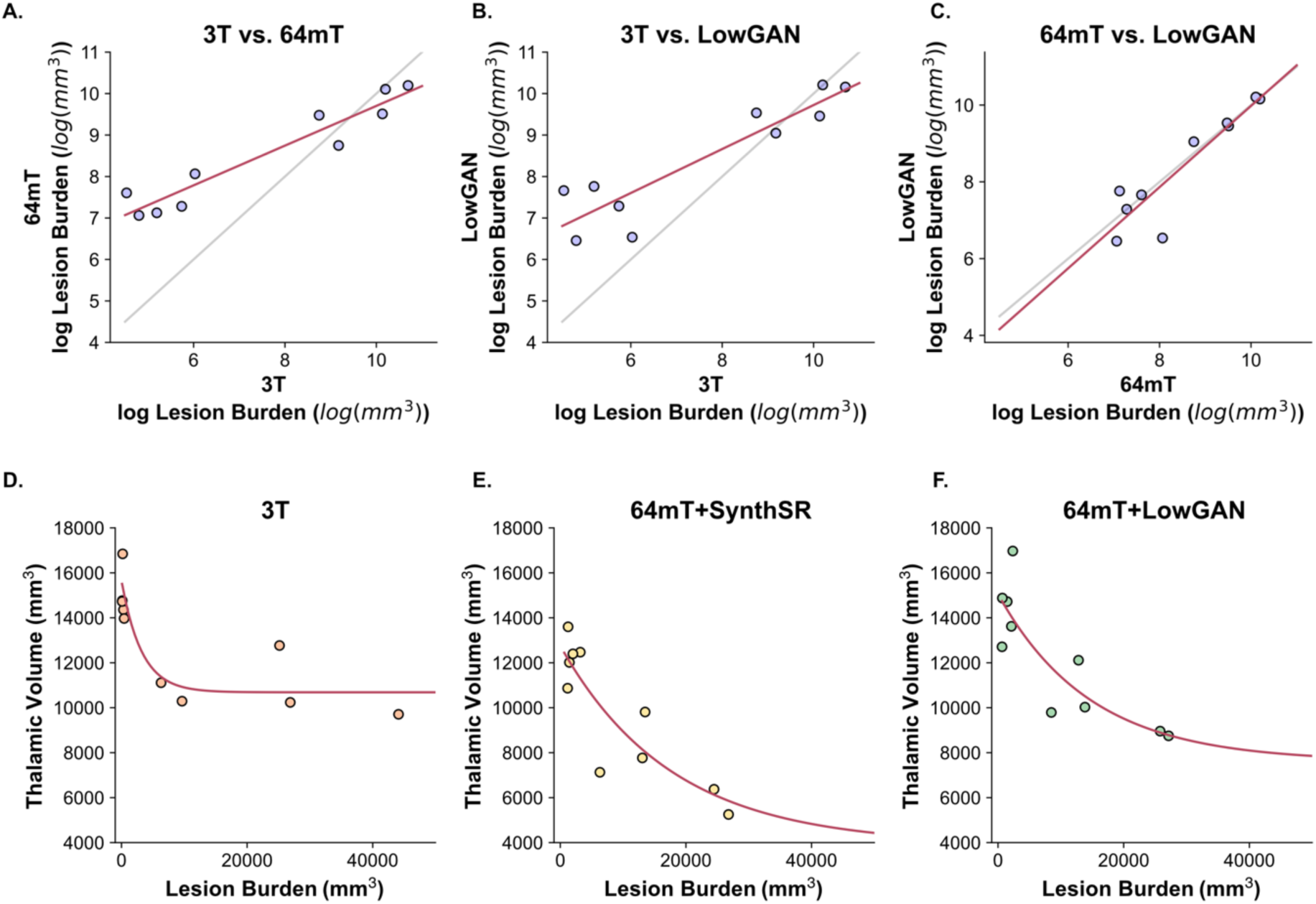
Lesion burden and its relationship with thalamic volume. Panels **A-C** show scatterplots of the logarithm of lesion burden estimated in test set cases for: **A.** high-field (3T) and low-field (64mT), **B.** high-field and LowGAN outputs, and **C.** low-field and LowGAN outputs. The black line represents perfect correspondence between both measurements and the red line represents the line-of-best-fit. Panels **D-F** show scatterplots of the relationship between measured thalamic volume and estimated lesion burden across test set cases for **D.** high-field (3T), **E.** low-field (64mT, with volumes estimated from SynthSR outputs), and **F.** LowGAN outputs. Red line represents the exponential line-of-best-fit. **a.u.** = arbitrary units.

## Discussion

Portable low-field MRI scanners present an opportunity to make neuroimaging more accessible in clinical and research settings where current availability is limited or absent. However, challenges related to image quality and resolution limit potential use cases. Our study addresses these challenges by developing a deep learning framework capable of synthesizing high-field quality images from low-field inputs, in this case applied to MS imaging.

### Quality Enhancement

Our results confirmed that the LowGAN synthetic high-field output images were not only visually superior but also quantitatively more correlated to high-field scans across T1w, T2w, and FLAIR sequences. The improvement was most pronounced in FLAIR, followed by T1w, and then T2w images. The quality enhancement observed in our study can potentially address the limitation of low-field scanners, making them more applicable for a broader range of diagnostic applications. Stage 1 outputs displayed the highest visual improvement, while stage 2 outputs, although visually less detailed, were necessary for segmentation. Either set of images could be used depending on the intended application. Deep learning techniques are being increasingly applied for low-field image enhancement^25^, and our results add to the promise of these approaches.

### Clinical Relevance

For MS, portable MRI could play a role in point-of-care diagnosis, screening, triage, or follow-up in cases where typical, particularly periventricular, white matter lesions of sufficient size are encountered^6^. Inevitably, a threshold exists for lesion detection and methods such as LowGAN to increase conspicuity are needed. Identifying brain atrophy has also become an important and complementary part of optimal clinical assessment in MS with the recognition that neurodegeneration occurs early in the disease course, may be partly independent of lesion activity, and better predicts disability and cognitive function^26^. As a result, many clinical trials of disease modifying therapies include atrophy as an outcome measure^27^. Thalamic volume in particular has emerged as a sensitive and robust marker of MS neurodegeneration and disease burden in clinical studies and drug trials^16,28–32^, potentially integrating direct and indirect injury across the brain via widespread cortical and subcortical connections. The ovoid shape of the thalamus and good contrast with adjacent CSF and white matter also benefit segmentation. LowGAN performed especially well in recapitulating 3T thalamic, ventricular and total cortical volumes, supporting a role for portable MRI in measuring brain atrophy. Lower cost, ease of use, and portability could all facilitate longitudinal imaging for clinical trials in MS, potentially improving robustness and reaching a broader range of the population including more severely affected patients^6^. Beyond MS, more accurate brain volume measurements, including for the hippocampus, are relevant for other etiologies of neurodegeneration or hydrocephalus that might be diagnosed or monitored with portable low-field MRI^5,33,34^.

### Comparison with Other Techniques

LowGAN compares favorably to SynthSR, the current state-of-the-art for portable MRI super resolution. LowGAN generates synthesized T1w, T2w and FLAIR images, whereas SynthSR currently only generates T1w images from T1w and T2w Hyperfine inputs. Of course, this comes with the additional requirement for FLAIR inputs. For brain morphometry, LowGAN outperformed SynthSR in most cases in producing brain region volumes approximating high-field scans. This may indicate that paired data better captures imaging features relevant to this transformation or to our specific scanners. Notably, we used standard sequences common across Hyperfine scanners. The addition of FLAIR contrast may also have improved fidelity. Our network could potentially be trained with fewer tissue contrasts to explore this possibility or provide more versatility. Adjacent slices could be incorporated into 2D training or a 3D architecture could be developed to make better use of volumetric information and obviate the need for two stage synthesis. Alternative architectures may have even better performance.

### Limitations and Future Directions

Limitations of our work include the specific focus on MS patients and the limited sample size. Optimally, future research will expand the availability of paired data for MS, other neurological conditions, and healthy participants. We expect lowGAN would perform similarly with other types of white matter lesions, but this requires testing and potentially additional paired data and training. Notably, our proposed framework is reversible, also enabling high-to-low field image translation. Low-field quality images synthesized in this way based on limited acquired paired data could expand available datasets for different disease processes with greater fidelity to true low-field characteristics, facilitating further software development, sequence optimization, and even synthetic clinical trials to evaluate different use cases^35^. For MS and more generally, future study with blinded readers will be needed to assess the true potential for improving brain lesion detection in portable MRI, and some combination of higher resolution imaging with longer scan times tailored to the pathology and clinical question, averaging of multiple full or partial acquisitions, and deep learning post-processing will likely prove optimal for this task. For volumetrics, additional data are needed to assess reliability and sensitivity for change over time.

## Conclusion

Synthesizing high-field-like images from low-field inputs using deep learning offers a means to enhance the quality and utility of low-field MRI in various clinical and research applications. For portable MRI in MS, LowGAN produces visually superior images, recovers clinically relevant brain volumes, increases lesion conspicuity, and preserves total white matter lesion burden. More generally, our paired MS dataset serves as a model for detecting both brain lesions and atrophy at low-field and our approach could be applied to other disease processes.

## Data Availability

The algorithm used in this manuscript will be made publicly available on GitHub upon publication of the manuscript. Data used in the manuscript will be made available upon reasonable request.

## Acknowledgments

We thank the team at Hyperfine, Inc. (Guilford, CT), particularly Jonathan Rothberg, PhD, and Megan Poorman PhD, and John Pitts, for technical assistance and the use of Hyperfine low-field MRI scanners. We thank the Penn Neuroradiology Research Core, including Lisa Desidero, Marisa Sanchez, Brian Dolan, Leeanne Lezotte, and Lauren Karpf, for assistance with patient recruitment and scanning. We also acknowledge the staff of the NINDS Neuroimmunology Clinic; Yeajin Song, MPS; Rose Cuento, CRNP; and the staff of the NIH NMR Center.

