## Supplementary for "Multi-contrast high-field quality image synthesis for portable low-field MRI using generative adversarial networks and paired data"

**Supplementary Table 1 - Fractional volumetry:** Comparison of intracranial volume (ICV) normalized volumes across structures. ICV is estimated from the 3T brain mask.

| Region | 3T vs SynthSR+64mT | | | 3T vs LowGAN+64mT | | | LowGAN+64mT vs SynthSR+64mT | | |
|---|---|---|---|---|---|---|---|---|---|
| | Δ Volume (mm$^3$)[a] | p-value[b] | Cohen's *d* | Δ Volume (mm$^3$)[a] | p-value[b] | Cohen's *d* | Δ Volume (mm$^3$)[a] | p-value[b] | Cohen's *d* |
| Left Thalamus | $1.1\times10^{-3}\pm1.0\times10^{-4}$ | **< 0.001** | 1.29 | $2.3\times10^{-4}\pm1.5\times10^{-4}$ | 0.544 | 0.26 | $9.0\times10^{-4}\pm1.4\times10^{-4}$ | **< 0.001** | 0.92 |
| Right Thalamus | $1.0\times10^{-3}\pm7.2\times10^{-5}$ | **< 0.001** | 1.26 | $3.2\times10^{-4}\pm1.0\times10^{-4}$ | **0.039** | 0.40 | $7.2\times10^{-4}\pm1.0\times10^{-4}$ | **< 0.001** | 0.84 |
| Left Lateral Ventricle | $-2.4\times10^{-3}\pm4.5\times10^{-4}$ | **0.001** | -0.44 | $-6.8\times10^{-6}\pm2.2\times10^{-4}$ | 0.999 | $-1.4\times10^{-3}$ | $-2.4\times10^{-3}\pm5.0\times10^{-4}$ | **0.002** | -0.46 |
| Right Lateral Ventricle | $-2.2\times10^{-3}\pm4.6\times10^{-4}$ | **0.002** | -0.47 | $8.7\times10^{-5}\pm1.9\times10^{-4}$ | 0.999 | 0.02 | $-2.3\times10^{-3}\pm5.1\times10^{-4}$ | **0.003** | -0.52 |
| Left Cerebral Cortex | $1.0\times10^{-2}\pm1.5\times10^{-3}$ | **< 0.001** | 1.36 | $-1.1\times10^{-4}\pm1.2\times10^{-3}$ | 0.999 | -0.01 | $1.0\times10^{-2}\pm1.6\times10^{-3}$ | **< 0.001** | 1.48 |
| Right Cerebral Cortex | $1.4\times10^{-2}\pm1.6\times10^{-3}$ | **< 0.001** | 1.81 | $-2.0\times10^{-3}\pm1.1\times10^{-3}$ | 0.364 | -0.27 | $1.6\times10^{-2}\pm1.5\times10^{-3}$ | **< 0.001** | 2.23 |
| Left Hippocampus | $2.7\times10^{-4}\pm7.2\times10^{-5}$ | **0.013** | 0.75 | $1.6\times10^{-4}\pm4.4\times10^{-5}$ | **0.016** | 0.70 | $1.1\times10^{-4}\pm9.6\times10^{-5}$ | 0.895 | 0.31 |
| Right Hippocampus | $3.1\times10^{-4}\pm9.6\times10^{-5}$ | **0.029** | 0.84 | $1.4\times10^{-4}\pm9.8\times10^{-5}$ | 0.595 | 0.45 | $1.7\times10^{-4}\pm1.5\times10^{-4}$ | 0.903 | 0.40 |
| Left Amygdala | $1.8\times10^{-4}\pm3.6\times10^{-5}$ | **0.002** | 1.66 | $5.9\times10^{-5}\pm3.8\times10^{-5}$ | 0.499 | 0.65 | $1.2\times10^{-4}\pm4.6\times10^{-5}$ | 0.080 | 0.96 |
| Right Amygdala | $1.8\times10^{-4}\pm3.4\times10^{-5}$ | **0.001** | 2.42 | $4.7\times10^{-5}\pm4.0\times10^{-5}$ | 0.863 | 0.42 | $1.3\times10^{-4}\pm3.6\times10^{-5}$ | **0.012** | 1.18 |
| Left White Matter | $3.5\times10^{-3}\pm2.3\times10^{-3}$ | 0.556 | 0.29 | $8.9\times10^{-4}\pm1.8\times10^{-3}$ | 0.999 | 0.07 | $2.6\times10^{-3}\pm1.5\times10^{-3}$ | 0.406 | 0.29 |
| Right White Matter | $7.8\times10^{-3}\pm2.5\times10^{-3}$ | **0.033** | 0.69 | $1.0\times10^{-3}\pm1.7\times10^{-3}$ | 0.999 | 0.08 | $6.8\times10^{-3}\pm1.7\times10^{-3}$ | **0.008** | 0.79 |
| Left Putamen | $2.0\times10^{-4}\pm1.1\times10^{-4}$ | 0.334 | 0.40 | $4.4\times10^{-4}\pm1.3\times10^{-4}$ | **0.026** | 1.15 | $-2.4\times10^{-4}\pm1.8\times10^{-4}$ | 0.657 | -0.53 |
| Right Putamen | $2.5\times10^{-4}\pm8.0\times10^{-5}$ | **0.041** | 0.52 | $5.9\times10^{-4}\pm1.2\times10^{-4}$ | **0.002** | 1.55 | $-3.5\times10^{-4}\pm1.0\times10^{-4}$ | **0.022** | -0.79 |
| Left Pallidum | $-9.4\times10^{-5}\pm2.5\times10^{-5}$ | **0.033** | -0.59 | $8.4\times10^{-5}\pm5.6\times10^{-5}$ | 0.530 | 0.57 | $-1.8\times10^{-4}\pm5.0\times10^{-5}$ | **0.018** | -1.32 |
| Right Pallidum | $-6.7\times10^{-5}\pm2.6\times10^{-5}$ | 0.099 | -0.42 | $6.5\times10^{-5}\pm4.5\times10^{-5}$ | 0.591 | 0.47 | $-1.3\times10^{-4}\pm4.4\times10^{-5}$ | **0.045** | -0.92 |
| Left Caudate | $1.4\times10^{-4}\pm5.1\times10^{-5}$ | 0.062 | 0.38 | $7.1\times10^{-6}\pm4.3\times10^{-5}$ | 0.999 | 0.02 | $1.4\times10^{-4}\pm5.6\times10^{-5}$ | 0.114 | 0.36 |
| Right Caudate | $2.1\times10^{-4}\pm4.8\times10^{-5}$ | **0.005** | 0.53 | $-2.9\times10^{-5}\pm5.5\times10^{-5}$ | 0.999 | -0.07 | $2.4\times10^{-4}\pm6.3\times10^{-5}$ | **0.013** | 0.59 |

[a]Volumes are computed as the residuals of a regression model created using the total intracranial volume between the first method and the second method. Δ Volume is calculated as the difference in mean volume of the region between the first method and second method. Mean +/- SE

[b]P-values are Bonferroni-corrected p-values for one-sample two-tailed t-tests assessing whether the Δ Volume is significantly different from zero

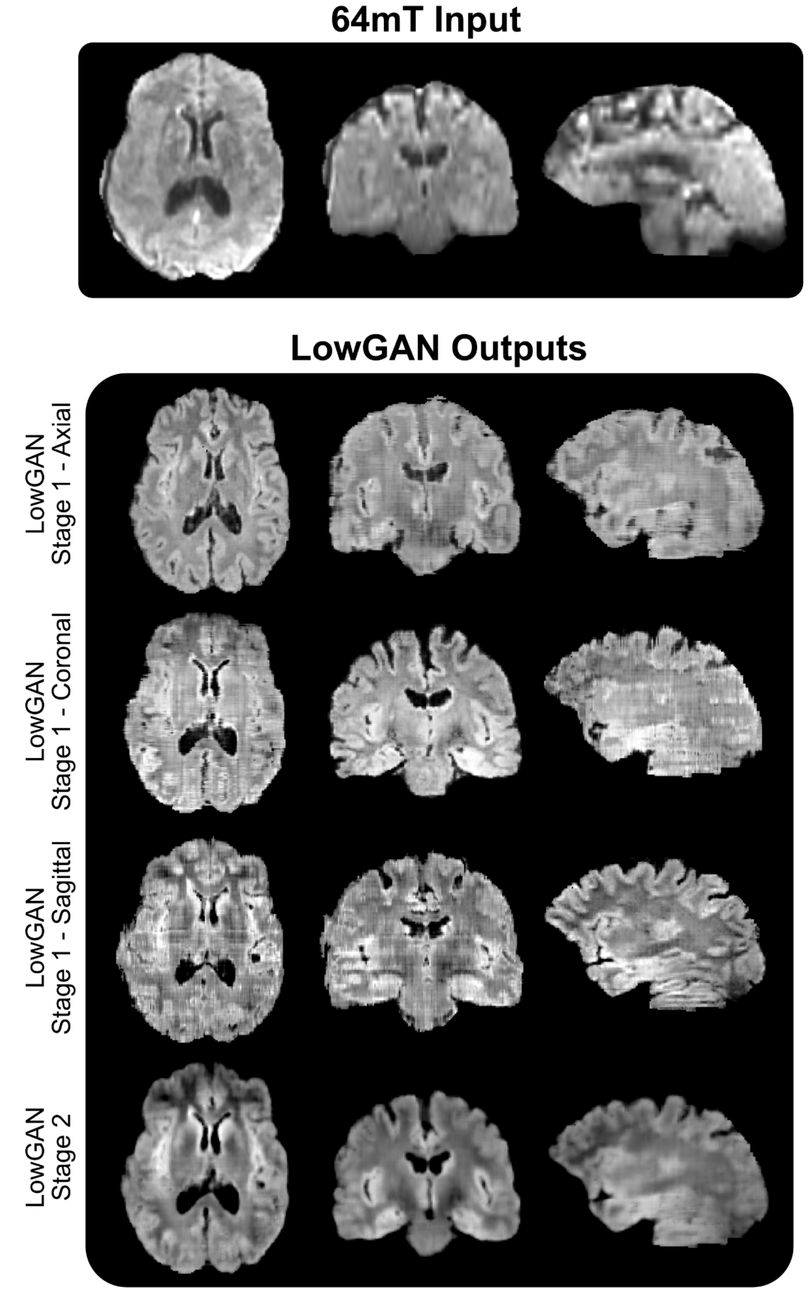


**Supplementary Figure 1 - Example FLAIR input slices and Stage 1 and 2 LowGAN outputs in orthogonal planes:** In Stage 1, 64mT axial, coronal, and sagittal input image stacks are processed with three separate corresponding orthogonal models. Here, output images are shown for the three orthogonal planes for each model. Note how Stage 1 outputs in planes orthogonal to the model plane (e.g. coronal and sagittal for the axial Stage 1) have horizontal or vertical lines across the slice. These are caused by intensity variations across slices in the model plane. Stage 2 generates volumes without these intensity variations, resulting in smoother volumes across the three orthogonal planes.

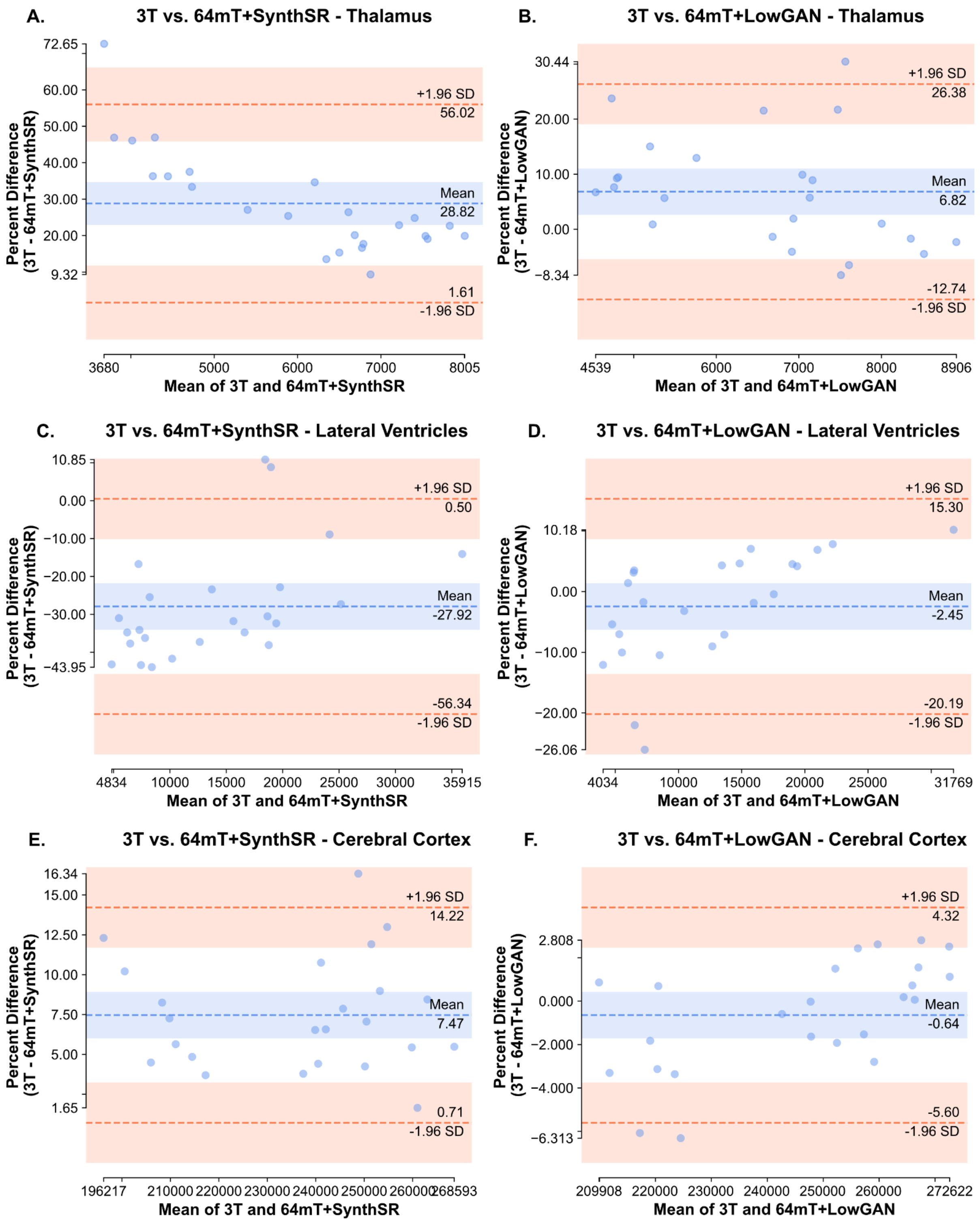


**Supplementary Figure 2:** Bland-Altman plots for comparisons between 3T and SynthSR volumes, and 3T and LowGAN volumes for the bilateral thalamus (**A** and **B**), lateral ventricles (**C** and **D**) and cortical gray matter (**E** and **F**).

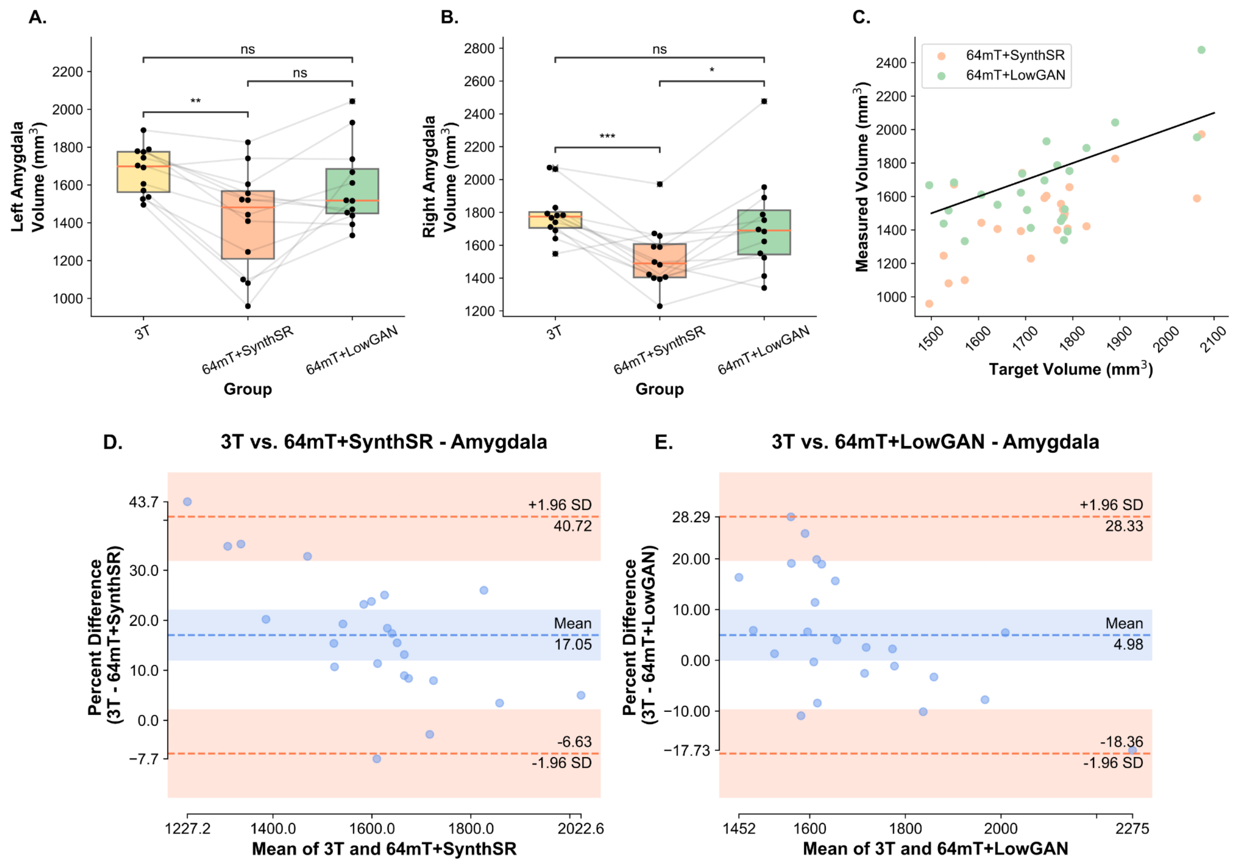


**Supplementary Figure 3 - Volumetry and Bland-Altman plots for the amygdala:** Panels **A** and **B** show the estimated left (**A**) and right (**B**) volumes for the amygdala, measured from SynthSeg segmentations. Lines connect the same participants across the boxplots for volumes measured at 3T, 64mT reconstructed with SynthSR (64mT+SynthSR) and 64mT reconstructed with LowGAN (LowGAN). Panel **C** has a scatterplot showing the relationship between the volume measured at high-field (target volume) and the volume measured in the SynthSR and LowGAN synthesized outputs (measured volume) for both left and right sided structures. The black line represents perfect correspondence between high-field and synthesized volumes. Panels **D** and **E** show Bland-Altman plots for comparisons between 3T and SynthSR volumes (**D**), and 3T and LowGAN volumes (**E**) for the left and right amygdala (both sides in the same plot).

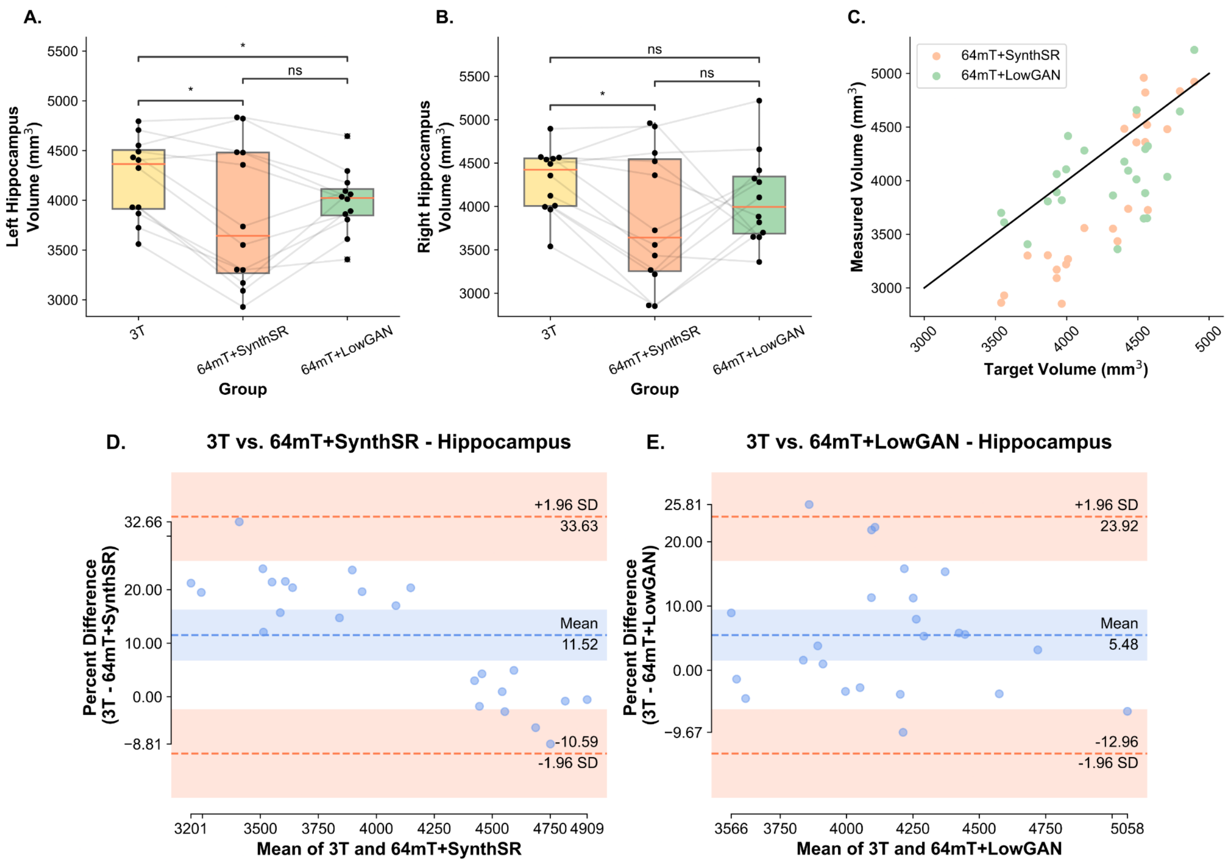


**Supplementary Figure 4 - Volumetry and Bland-Altman plots for the hippocampus:** Panels **A** and **B** show the estimated left (**A**) and right (**B**) volumes for the hippocampus, measured from SynthSeg segmentations. Lines connect the same participants across the boxplots for volumes measured at 3T, 64mT reconstructed with SynthSR (64mT+SynthSR) and 64mT reconstructed with LowGAN (LowGAN). Panel **C** has a scatterplot showing the relationship between the volume measured at high-field (target volume) and the volume measured in the SynthSR and LowGAN synthesized outputs (measured volume) for both left and right sided structures. The black line represents perfect correspondence between high-field and synthesized volumes. Panels **D** and **E** show Bland-Altman plots for comparisons between 3T and SynthSR volumes (**D**), and 3T and LowGAN volumes (**E**) for the left and right hippocampus (both sides in the same plot).

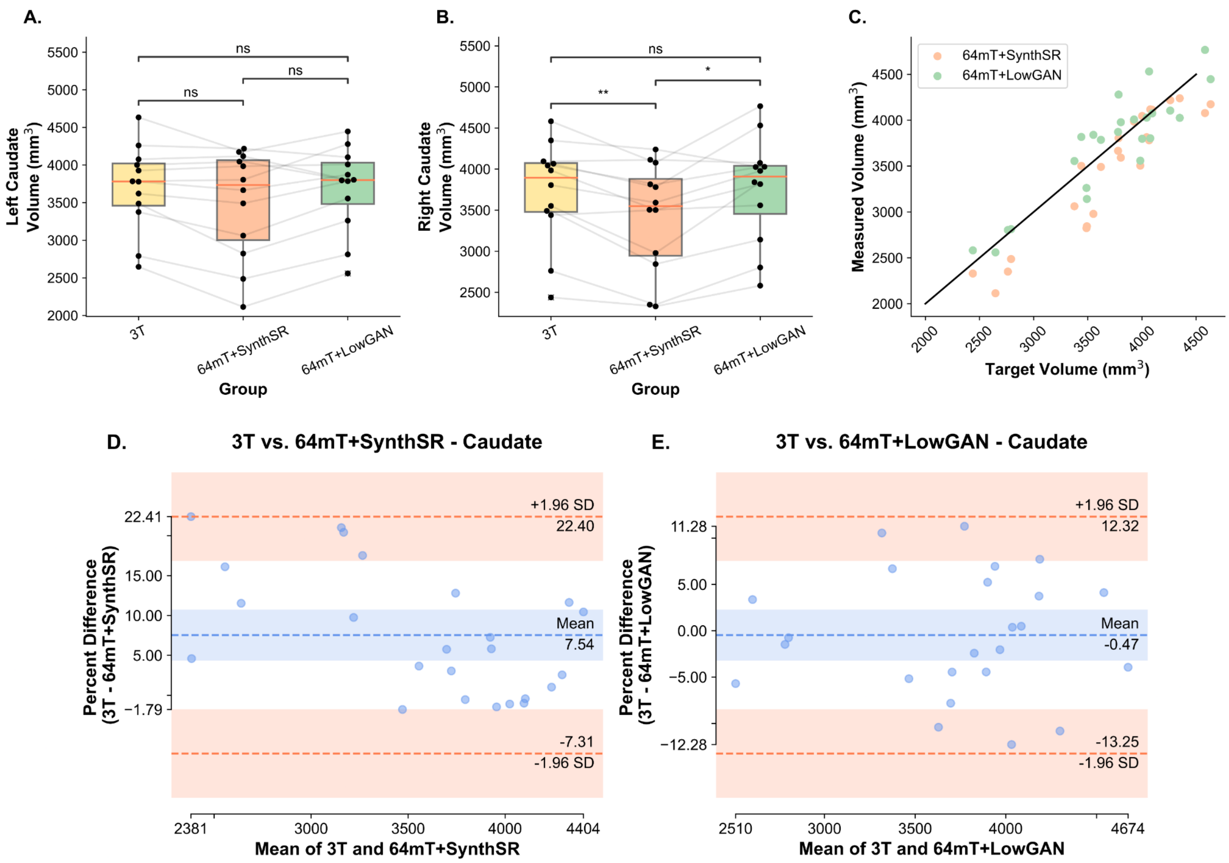


**Supplementary Figure 5 - Volumetry and Bland-Altman plots for the caudate:** Panels **A** and **B** show the estimated left (**A**) and right (**B**) volumes for the caudate, measured from SynthSeg segmentations. Lines connect the same participants across the boxplots for volumes measured at 3T, 64mT reconstructed with SynthSR (64mT+SynthSR) and 64mT reconstructed with LowGAN (LowGAN). Panel **C.** has a scatterplot showing the relationship between the volume measured at high-field (target volume) and the volume measured in the SynthSR and LowGAN synthesized outputs (measured volume) for both left and right sided structures. The black line represents perfect correspondence between high-field and synthesized volumes. Panels **D** and **E** show Bland-Altman plots for comparisons between 3T and SynthSR volumes (**D**), and 3T and LowGAN volumes (**E**) for the left and right caudate (both sides in the same plot).

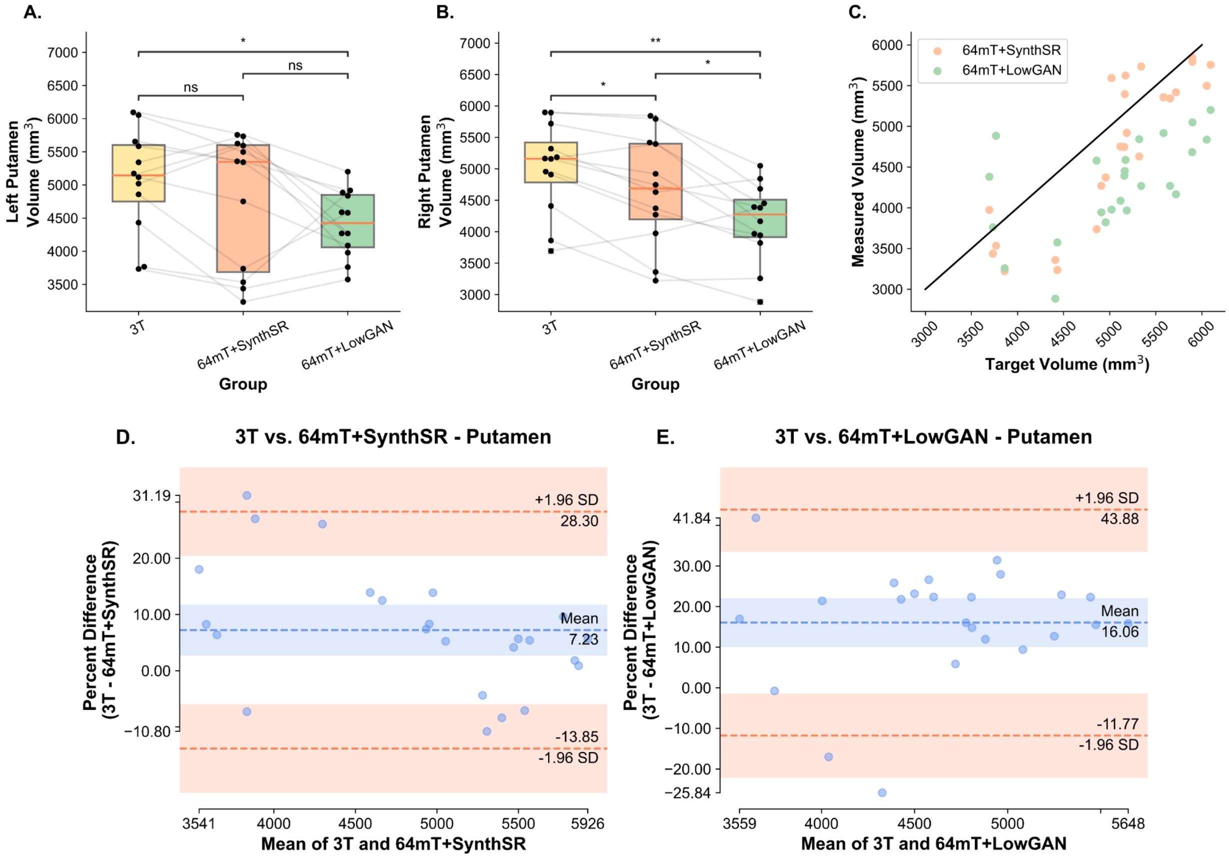


**Supplementary Figure 6 - Volumetry and Bland-Altman plots for the putamen:** Panels **A** and **B** show the estimated left (**A**) and right (**B**) volumes for the putamen, measured from SynthSeg segmentations. Lines connect the same participants across the boxplots for volumes measured at 3T, 64mT reconstructed with SynthSR (64mT+SynthSR) and 64mT reconstructed with LowGAN (LowGAN). Panel **C** has a scatterplot showing the relationship between the volume measured at high-field (target volume) and the volume measured in the SynthSR and LowGAN synthesized outputs (measured volume) for both left and right sided structures. The black line represents perfect correspondence between high-field and synthesized volumes. Panels **D** and **E** show Bland-Altman plots for comparisons between 3T and SynthSR volumes (**D**), and 3T and LowGAN volumes (**E**) for the left and right putamen (both sides in the same plot).

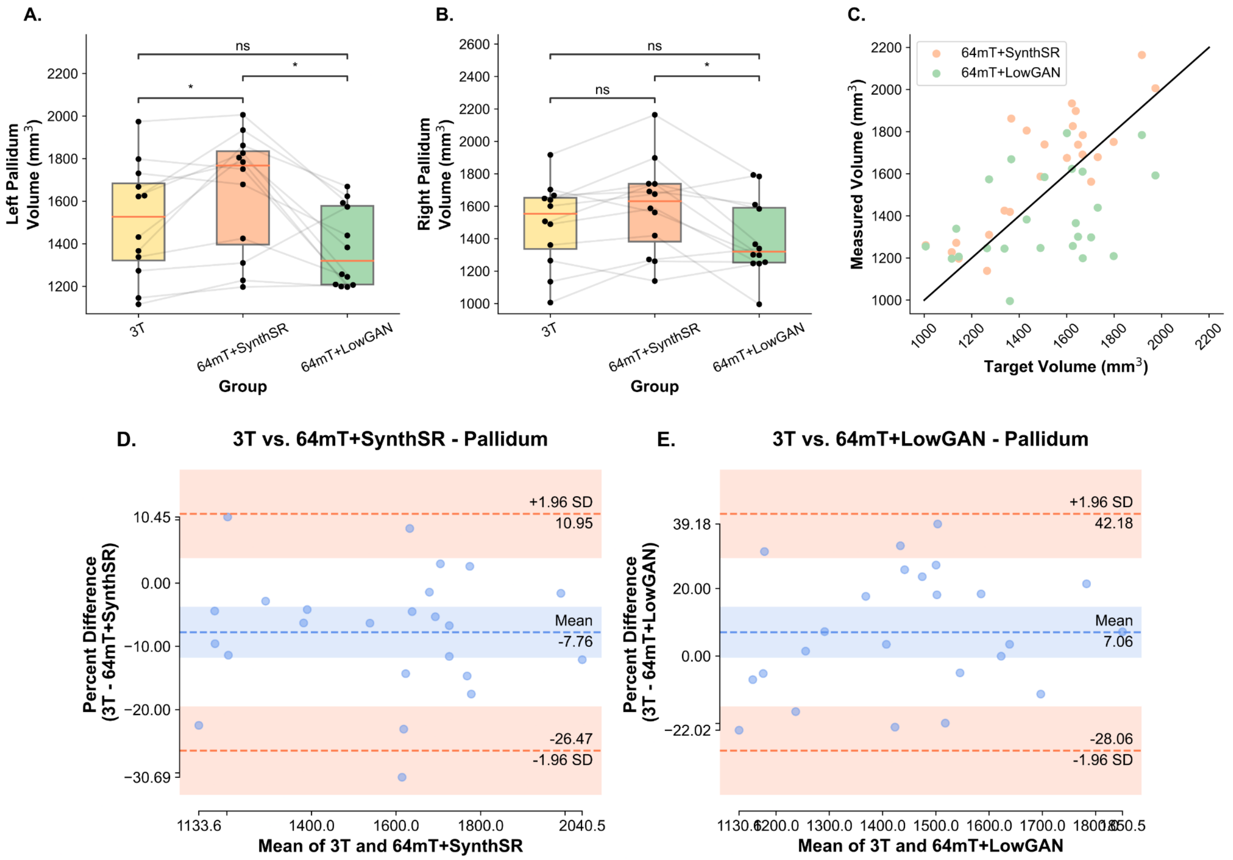


**Supplementary Figure 7 - Volumetry and Bland-Altman plots for the pallidum:** Panels **A** and **B** show the estimated left (**A**) and right (**B**) volumes for the pallidum, measured from SynthSeg segmentations. Lines connect the same participants across the boxplots for volumes measured at 3T, 64mT reconstructed with SynthSR (64mT+SynthSR) and 64mT reconstructed with LowGAN (LowGAN). Panel **C** has a scatterplot showing the relationship between the volume measured at high-field (target volume) and the volume measured in the SynthSR and LowGAN synthesized outputs (measured volume) for both left and right sided structures. The black line represents perfect correspondence between high-field and synthesized volumes. Panels **D** and **E** show Bland-Altman plots for comparisons between 3T and SynthSR volumes (**D**), and 3T and LowGAN volumes (**E**) for the left and right pallidum (both sides in the same plot).

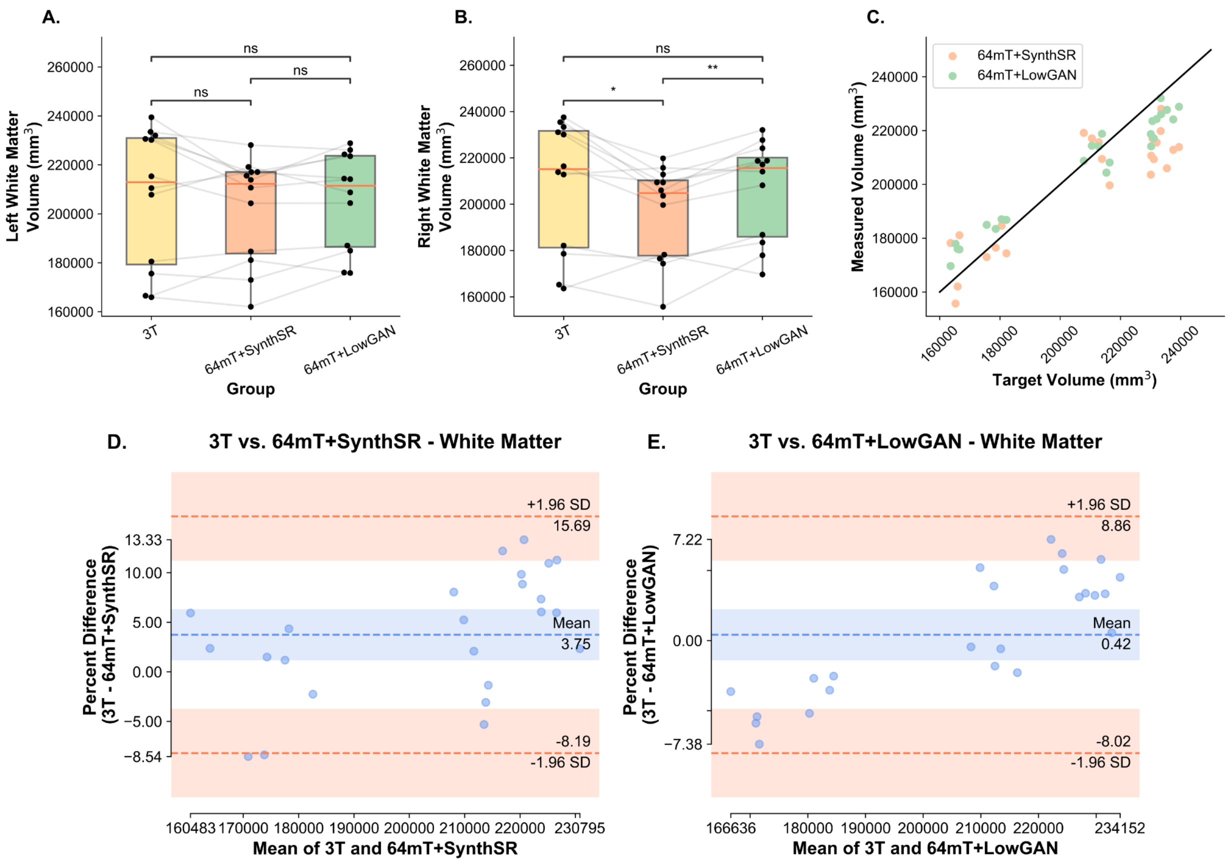


**Supplementary Figure 8 - Volumetry and Bland-Altman plots for white matter:** Panels **A** and **B** show the estimated left (**A**) and right (**B**) volumes for the white matter, measured from SynthSeg segmentations. Lines connect the same participants across the boxplots for volumes measured at 3T, 64mT reconstructed with SynthSR (64mT+SynthSR) and 64mT reconstructed with LowGAN (LowGAN). Panel **C** has a scatterplot showing the relationship between the volume measured at high-field (target volume) and the volume measured in the SynthSR and LowGAN synthesized outputs (measured volume) for both left and right sided structures. The black line represents perfect correspondence between high-field and synthesized volumes. Panels **D** and **E** show Bland-Altman plots for comparisons between 3T and SynthSR volumes (**D**), and 3T and LowGAN volumes (**E**) for the left and right white matter (both sides in the same plot).
